# Quantifying MS Progression in the Era of Highly Effective Therapy: Trial Design Implications

**DOI:** 10.64898/2026.05.06.26352552

**Authors:** Arnaud Gaudry, Gian-Andrea Thanei, H.-Christian von Büdingen, Stephen Krieger, James Overell, Maria Pia Sormani, Ulrike Bonati, Marcelo Boareto

**Author notes:** **Corresponding author:** Arnaud Gaudry, PhD, Roche Pharma Product Development (PD), F. Hoffmann-La Roche Ltd., Grenzacherstrasse 124, 4070 Basel, Switzerland. These authors contributed equally (Ulrike Bonati, Marcelo Boareto).

## Abstract

**Importance:** In multiple sclerosis (MS), high-efficacy disease-modifying therapies (HEDMTs) effectively control relapse-associated worsening (RAW), but progression independent of relapse activity (PIRA) remains inadequately addressed. As HEDMTs become the standard of care, developing new therapies that target this residual progression is a critical unmet need.

**Objective:** This study quantifies disability progression in MS patients treated with ocrelizumab to evaluate how confirmed EDSS disability progression (EDSS-CDP) would perform as an endpoint in future trials using HEDMT as comparators.

**Design:** Retrospective longitudinal cohort study.

**Setting:** Pooled dataset from four multicenter phase III and IV clinical trials.

**Participants:** 1,859 people with (pw) relapsing MS (RMS), primary progressive MS (PPMS), and secondary progressive MS (SPMS) who were treated with ocrelizumab within the OPERA I/II, ORATORIO, and CONSONANCE trials.

**Intervention:** Ocrelizumab.

**Main Outcomes and Measures:** We developed a hierarchical Bayesian model to analyze longitudinal EDSS trajectories using two components: an offset effect, used to capture changes occurring rapidly after treatment onset, followed by a steady, long term linear progression over time. We used this model to simulate future clinical trial scenarios, assuming different drug effects on the offset and the long term linear progression.

**Results:** Our model accurately describes longitudinal EDSS changes and the risk of EDSS-CDP in ocrelizumab-treated subjects. Disability improvement (offset effect) was most prominent in pwRMS, while pwPPMS exhibited the highest long-term progression rates. Baseline T1 gadolinium-enhancing lesions were associated with a greater initial benefit. Simulations of typical phase III trials suggest that the hazard ratio on the EDSS-CDP endpoint is mostly influenced by the magnitude of the offset effect rather than the impact on long-term linear progression.

**Conclusions and Relevance:** We attribute the disability improvement observed shortly after treatment onset to resolving focal inflammation, and the long-term steady progression rate to disease mechanisms not fully addressed by ocrelizumab. Our simulation results show that within the current trial paradigm, which uses EDSS-CDP as a measure of disability progression, the ability of a treatment to induce an initial improvement is the primary determinant of success. These results emphasize the urgent need for both innovative clinical trial designs and more sensitive endpoints to adequately assess the next generation of MS therapies targeting gradual disability progression.

*Key Points:* Question
Will the standard multiple sclerosis disability progression endpoint, confirmed EDSS disability progression (EDSS-CDP), prove to be an accurate measure of the efficacy of new therapies addressing long-term progression when compared against high-efficacy treatments (HET)? Findings
In this modeling study of 10-year ocrelizumab data, observed changes in EDSS were characterized by an early improvement followed by a linear long-term worsening. EDSS-CDP was shown to be highly sensitive to initial improvement. Since this phenomenon strongly influences the overall treatment effect, trials that use ocrelizumab, or similar HET as a comparator may fail to identify novel treatments designed to further slow long-term progression. Meaning
Current trial designs may be inadequate for evaluating next-generation MS therapies, necessitating the development of better metrics to capture treatment effects on gradual progression.

## Introduction

Multiple sclerosis (MS) is a chronic, immune-mediated, disabling disease affecting around 2.9 million people worldwide.^1^ The clinical course of MS can be described using three different phenotypic descriptors:^2^ relapsing (RMS), secondary progressive (SPMS), and primary progressive (PPMS). Current evidence suggests that these phenotypes belong to the same disease spectrum, and that the observed distinctions can be attributed to differences in the degree to which the various underlying pathological mechanisms manifest clinically.^3–6^ The clinical manifestation of disability worsening is observed through two distinct mechanisms: incomplete recovery following a relapse due to focal inflammation, called relapse-associated worsening (RAW), and progression independent of relapse activity (PIRA) in which disability worsening occurs as a result of progressive central nervous system (CNS) disease burden due to gradual pathological processes, including compartmentalised CNS inflammation and neurodegeneration.^7^

The most widely used MS disability measure is the Expanded Disability Status Scale (EDSS). The EDSS is a composite scale ranging from 0 to 10 in incremental steps of 0.5 points, with lower scores indicating less disability and higher scores indicating greater disability.^8^ Worsening of the EDSS score, which is confirmed after a specified time (for example 12 or 24 weeks), is referred to as confirmed disability progression (CDP). A typical EDSS-CDP24 definition is an increase in EDSS of at least 1.0 point if the EDSS is inferior to 6, or 0.5 points if EDSS is 6 or greater, confirmed after 24 weeks. EDSS-CDP is used in MS clinical trials as a time-to-event endpoint to evaluate the effect of a therapy on the risk of disability progression. In such trials, subjects are typically randomized to a control or a treatment arm, and their individual on-trial EDSS scores are compared to their EDSS score at baseline to derive the time to first EDSS-CDP event. The statistical relevance of an eventual treatment effect is then typically assessed using a Cox proportional hazards (CoxPH) regression model to estimate the hazard ratio (HR), *i*.*e*. the relative risk of experiencing a CDP event in the treatment arm compared to the control arm at any given point in time.

A variety of disease-modifying therapies (DMTs) are available to MS patients and clinicians. Ocrelizumab, an anti-CD20 monoclonal antibody, demonstrates high efficacy in suppressing disease activity—measured by MRI lesions and relapses—as well as a significant reduction in disability progression. In OPERA I and II, the two pivotal phase III trials comparing ocrelizumab with interferon beta-1a in RMS patients, ocrelizumab showed almost complete suppression of new T1 Gd-enhancing lesions; 94% and 95% over a 96-week period in OPERA I and OPERA II compared to interferon (IFN) beta-1a, respectively, and new or enlarging hyperintense T2 lesions (77% and 83% respectively).^9^ Clinically, this translated into an annualized relapse rate (ARR) of 0.16 in both trials, representing a reduction of 46% and 47%, respectively, compared to interferon beta-1a.^9^ Regarding ocrelizumab’s effect on disability progression, both OPERA trials revealed a significant reduction, with a HR of 0.57 (95% CI, 0.34-0.95) and 0.63 (0.40-0.98) on EDSS-CDP24 against IFN beta-1a, for OPERA I and II respectively.^9^ In the ORATORIO phase III trial, comparing ocrelizumab with placebo in PPMS, a significant effect on EDSS-CDP24 was found with a HR of 0.75 (0.58–0.98).^10^

Together, these findings demonstrate a profound effect of ocrelizumab on the focal inflammatory component of MS, which translates into a significant effect on ARR. They also demonstrate a highly significant effect of ocrelizumab on disability progression. Despite this, disability progression continues to occur in pwMS treated with ocrelizumab and represents a substantial remaining unmet need. Current evidence shows that the residual progression is mainly attributable to PIRA, which occurs as a consequence of multiple pathological mechanisms unaddressed by current high-efficacy DMTs (HEDMTs).^7,11,12^ With the rising availability of HEDMTs and their establishment as the new standard of care, new drug candidates are likely to be tested against or as addition to such treatments in randomized trials, and judged clinically on their effect on measures of disability progression. In this new context, a deeper understanding of the disability progression that occurs in pwMS treated with HEDMTs and its measurement is needed.

This study presents a mathematical model to quantify long-term disability progression in pwMS treated with ocrelizumab, with the objective to inform the design of future clinical trials targeting MS progression. Using longitudinal EDSS change data from different clinical trials, we characterize and distinguish between short-term and long-term disability dynamics following treatment initiation and assess their association with baseline characteristics. We then establish a relationship between the model parameters and the cumulative risk of confirmed disability progression. This model is used to simulate treatment scenarios to estimate the expected effect size on EDSS-CDP, expressed as a HR, for novel therapies targeting disability progression when compared with HEDMTs.

## Methods

### Study Population, Registrations, and Patient Consents

Overall, 763 ocrelizumab treated pwRMS (383 from OPERA I [NCT01247324] and 380 from OPERA II [NCT01412333]); 743 ocrelizumab treated pwPPMS (445 from ORATORIO [NCT01194570]; and 298 from CONSONANCE [NCT03523858]); and 353 ocrelizumab treated pwSPMS (from CONSONANCE); were included. All participants had no missing covariates and at least 4 EDSS assessments.^9,10,13^ This resulted in the inclusion of 1,859 subjects, and a total of 37,478 EDSS assessments. The median follow-up durations per study were 9.7, 9.3, 9.7, and 3.4 years for OPERA I, OPERA II, ORATORIO, and CONSONANCE, respectively. Demographics and baseline characteristics are available in Supplementary Table 1.

### Ethics

The current study is a secondary analysis of anonymised patient data from clinical trials fully approved by institutional review boards and local regulatory authorities. All patients provided written informed consent.

### Mathematical Model

Our analysis of disability progression in pwMS treated with ocrelizumab involved specifying a hierarchical Bayesian model. Based on the empirical observations of change from baseline in EDSS, we modeled it using a non-linear function of time composed of two additive components. The first component is an offset effect, characterized by two parameters, *maximum offset effect* and *steepness* (which can be interpreted as the time needed to reach half of the *maximum offset effect*). The second component is a linear progression, characterized by a single *progression rate* parameter (**Figure 1A**). The *maximum offset effect* and *progression rate* parameters were modeled hierarchically to account for subject-level variability, while the steepness of the offset effect was modeled as a global parameter.^14^

**Figure 1:**
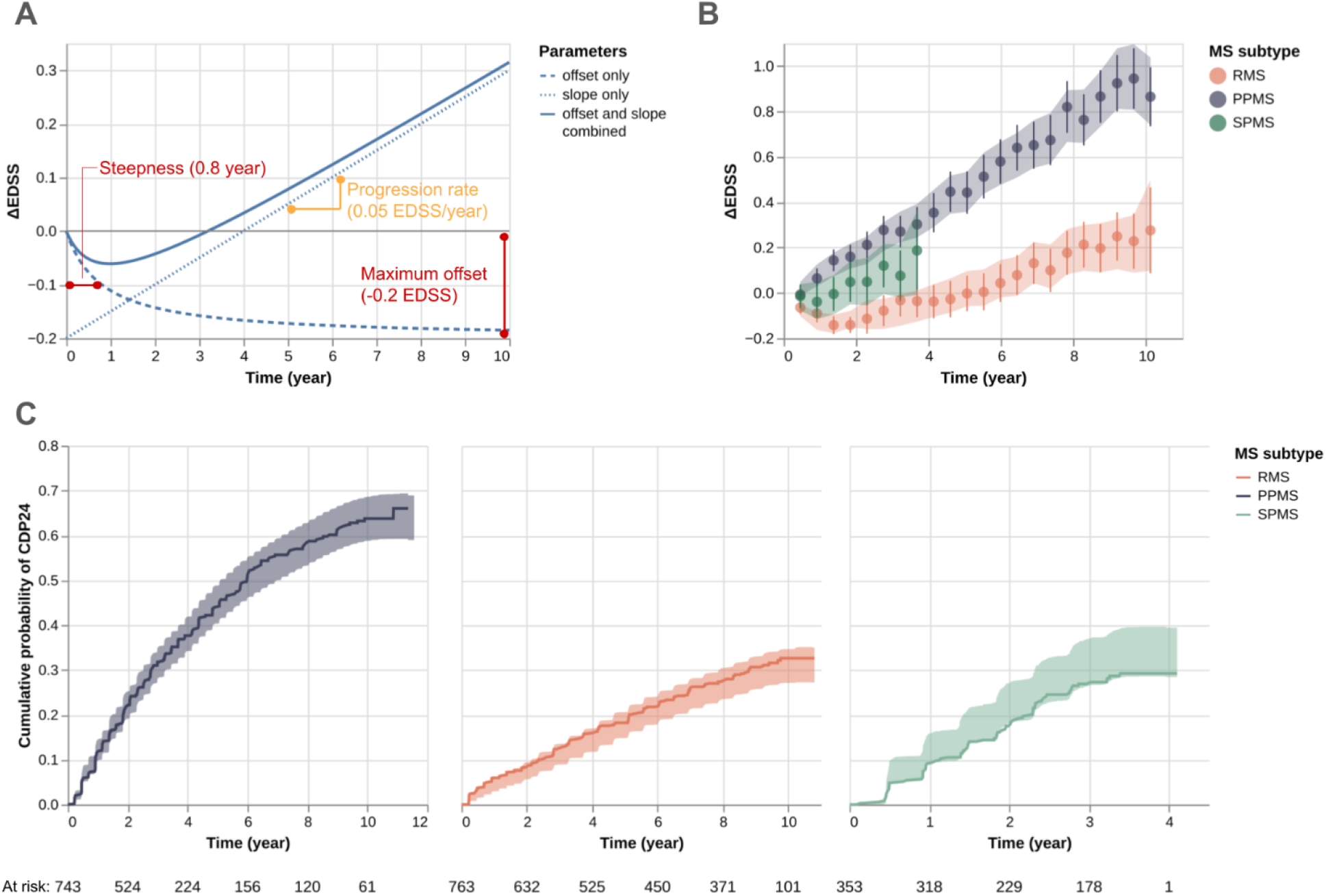
Model description, EDSS change from baseline and EDSS-CDP24. A) Visualization of the two model components, offset effect and linear, independently and combined. The impact of the two parameters defining the offset effect, *steepness* and *maximum offset*, is depicted in red, with in parentheses the value used for this visualization. The impact of the parameter driving the linear progression, the *progression rate*, is depicted in orange. B) Observed EDSS change from baseline (ΔEDSS, dots represent the mean and vertical lines the 95% CI), and model posterior predictive 95% CI (shaded area), per MS subtype. C) Cumulative probability of EDSS-CDP24, observed (survival curve, bold solid line) and predicted from the model (95% CI, shaded area). The numbers at risk are for the observed data. RMS: relapsing MS, PPMS: primary progressive MS, SPMS: secondary progressive MS.

Patient characteristics at baseline were evaluated as potential predictive covariates for both the *maximum offset effect* and the *progression rate*. The following covariates were included: MS subtype (PPMS, or SPMS), age, BMI, T2 lesions volume, years since disease onset (first symptoms of MS), EDSS, sex and presence/absence of T1 Gd+ lesions. RMS is not included as a covariate since it is considered the “default” MS subtype in the model, while PP and SPMS were integrated as covariates to evaluate how a *typical* pwPPMS or pwSPMS differ from a typical pwRMS. To facilitate the interpretation and comparison of regression coefficients, continuous covariates (age, BMI, T2 lesions volume, years since disease onset, and EDSS) were z-standardized. A linear relationship between the covariates was tested as predictive factors. Nonlinear relationships between the covariates were not addressed in this analysis.

To implement our model, we used the probabilistic programming language Stan, which enables us to perform full Bayesian inference.^15^ Specifically, we used Hamiltonian Monte Carlo, a Markov chain Monte Carlo method, to generate samples from the posterior distribution. More details about the model are available in Supplementary Information, and the posterior distribution of model key-parameters can be found in Supplementary Table 2.

### Clinical Trials Simulations

Simulations were performed using a mathematical model similar to the one used for the analysis of disability progression, but without any covariate effect on the *maximum offset effect* and *progression rate* parameters. For each clinical trial simulation, the fixed slope and fixed offset were set to specified values, for each treatment arm (control and treatment). For each subject (500 per treatment arm) random effects for both the slope and the offset were drawn from a Laplace distribution. Random effects distribution location and scale parameters, assumed identical for both arms, were drawn from the posterior distribution previously estimated from the data (fitted model). Similarly, the steepness of the offset effect and the residual error were drawn for the posterior distribution of the fitted model. Then, these parameters were used to generate EDSS-change-from-baseline values at each time point (every 12 weeks, until week 144 post-baseline) for each subject.

Since baseline EDSS values are needed to derive EDSS-CDP events and are not simulated by the model, these values were drawn for each simulation from the pool of pwPPMS included in the study cohort, with replacement. The resulting dataset, comprising baseline EDSS and EDSS-change from baseline, was used to derive, for each patient, the time to first EDSS-CDP24 event. Then, as typically done in clinical trials, a CoxPH regression model was fitted to obtain the HR between the treatment arms and the corresponding *p*-value. The core assumption of this model is that the hazards are proportional, meaning the ratio of the risk of an event between the two arms remains constant over time. The primary output, the hazard ratio, represents the relative risk of experiencing a CDP event in the treatment arm compared to the control arm at any given point in time. An HR<1.0 indicates a reduction in risk for the treatment arm, with an HR of 0.7, e.g. signifying a 30% lower risk of confirmed progression compared to the control. Conversely, an HR>1.0 would imply an increased risk, while an HR=1.0 suggests no difference between the arms. This process was repeated 1,000 times for each comparator slope-treatment effect pair to obtain the average HR and the estimated power for this pair.

### Software

Data processing, figure generation, simulations and statistical analysis were performed in *Python* v3.13, and *cmdstanpy* v1.2.4 was used for inference.

### Source Code

The source code used to run this analysis can be found at: https://github.com/ArnaudGaudry/ms-edss-modeling.

### Data Availability

Qualified researchers may request access to individual patient-level clinical data through a data request platform. At the time of writing, this request platform is Vivli (vivli.org/ourmember/roche/). Up-to-date details on Roche”s Global Policy on the Sharing of Clinical Information and how to request access to related clinical study documents can be found at go.roche.com/data_sharing. Anonymized records for individual patients across more than one data source external to Roche cannot, and should not, be linked because of a potential increase in risk of patient re-identification.

## Results

### 1. EDSS Change Model and link with EDSS-CDP

We characterized the average EDSS trajectory for ocrelizumab treated individuals into two components; an initial decrease or stabilization of EDSS occurring in the first months following treatment start, and an underlying gradual disability progression that becomes evident in the months/years that follow the initial response. Mathematically, we captured the initial decrease or stabilization of EDSS using a non-linear *offset effect*, and the gradual disability progression using a *linear* function. The offset effect is defined by two parameters, *steepness* and *maximum offset*, while the linear function is defined by a single slope parameter, corresponding to the long term *progression rate*.

Using different combinations of steepness and progression rate parameters, this model allows to describe different patterns of progression (**Fig. 1A**), and the observed EDSS mean change from baseline for the different MS subtypes (**Fig. 1B**). To quantify and account for interindividual variability among PwMS, we included subject-specific random effects in the model, for both offset effect and progression rate.

We could demonstrate that this structure efficiently accounts for both the mean EDSS change from baseline and EDSS-CDP (**Fig. 1B,C**). Specifically, it captures the mean change from baseline at the group level (**Fig. 1B**, shaded area), as well as the underlying distribution and spread of EDSS change from baseline data (**Fig. S1**). In addition, the model is able to reproduce EDSS-CDP24 cumulative probability (**Fig. 1C**), providing a framework for evaluating the impact of different EDSS trajectories on this endpoint.

### 2. Differences Between the MS subtypes and Effect of T1-Gd Enhancing Lesions at Baseline

We used the model to quantify differences in disease progression between the different MS subtypes, and the impact of 7 other baseline covariates representing patient demographics or disease-specific information **(Fig. 2** and **Fig. S2**). The effect of these covariates was evaluated for both the progression rate and the offset effect. Regarding the MS subtypes” impact, the progression rate was highest for PPMS, intermediate for SPMS, and lowest for RMS (**Fig. 2A**). Regarding the offset effect, the opposite was observed, with a strong offset effect observed for RMS, intermediate for SPMS, and lowest for PPMS (**Fig. 2B**). A cautious interpretation of the SPMS parameters is however required, given the larger credible intervals for this MS subtype, caused by the smaller sample size and lack of long term follow-up data (> 5 years) for this group.

**Figure 2:**
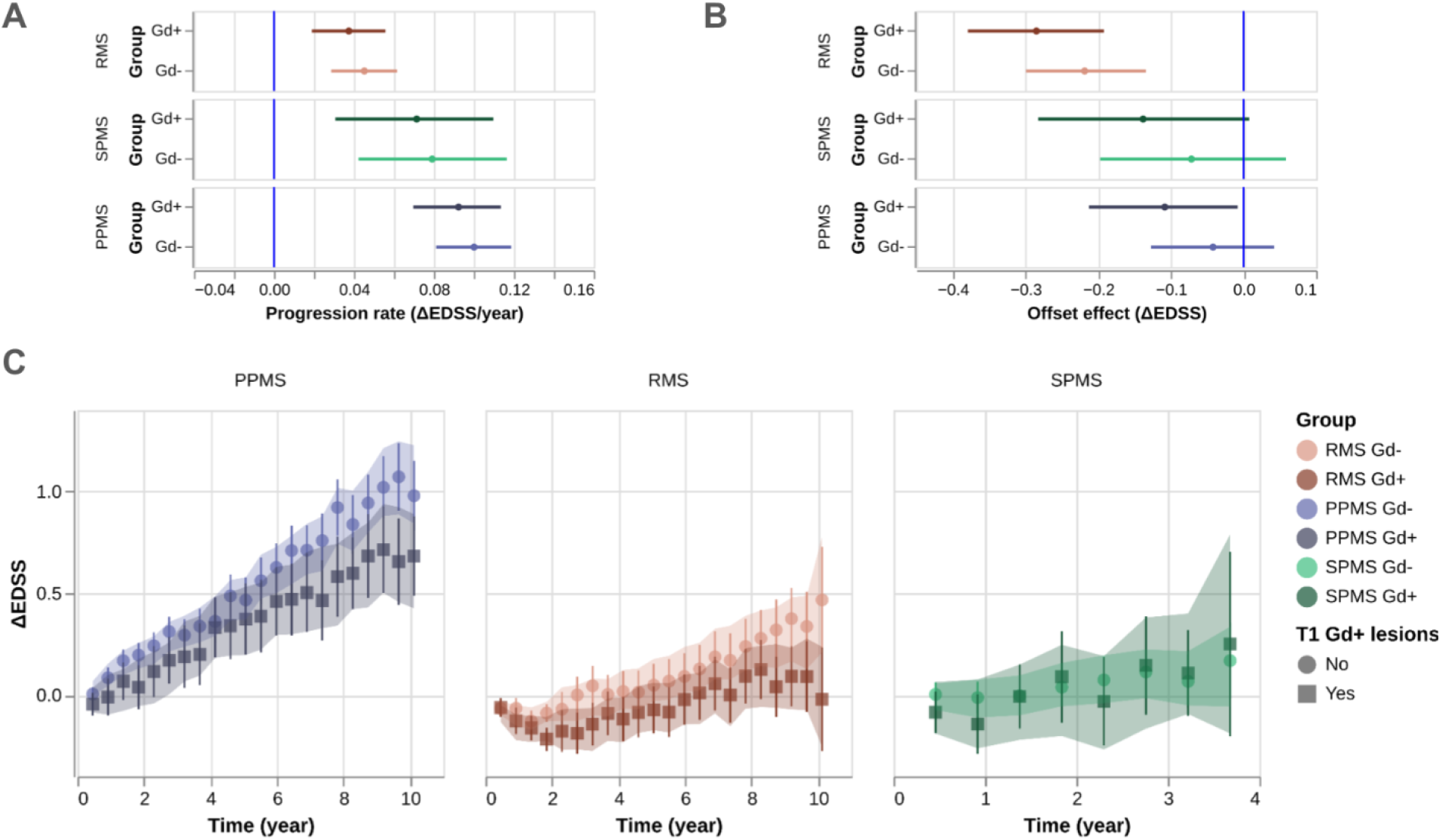
Progression rate and offset effect parameters values for different groups, as well as EDSS change from baseline for these groups. A) Progression rate parameters for a typical patient from each MS subtype with (Gd +) and without (Gd -) T1-Gd enhancing lesions at treatment initiation (mean and 95% credible interval). B) Offset effect parameters for a typical patient from the same groups as panel A (mean and 95% credible interval). C) Observed EDSS change from baseline (ΔEDSS, points and 95% CI), and model posterior predictive 95% CI (shaded area), for each group of panels A and B. For A and B, the vertical blue line represents a null effect.

It is important to note that the minimal dropout rate observed at 2 years (12% over all MS subtypes) suggests that the initial improvement after baseline is not influenced by dropout effects (**Fig. S3A**). This low dropout rate enables a robust estimation of the parameters for the offset effect, as this effect is primarily determined by the differences between baseline and initial follow-up visits. For the ORATORIO and OPERA trials, the dropout rate increased to approximately 25% at 5 years and to 47% and 41% at 9 years, respectively.

When studying the impact of baseline covariates other than the MS subtype on the progression rate and the offset effect, the presence of T1 Gd+ lesions at baseline, a marker of disease activity, seemed to be associated with a larger offset effect and a lower rate of long-term progression (**Fig. 2**). This effect is particularly visible for the PPMS and RMS subtypes (**Fig. 2C**). The impact of other covariates on the offset and slope parameters is presented in **Fig. S3**. At a 95% CI threshold, age and male sex were associated with faster progression rates, while a higher EDSS at baseline was associated with a more significant offset effect.

### 3. Clinical Trial Simulations and Impact on EDSS-CDP

We then investigated, through simulations, three different clinical trial scenarios for drugs targeting the residual progression observed in pwMS treated with ocrelizumab (**Fig. 3)**.

**Figure 3:**
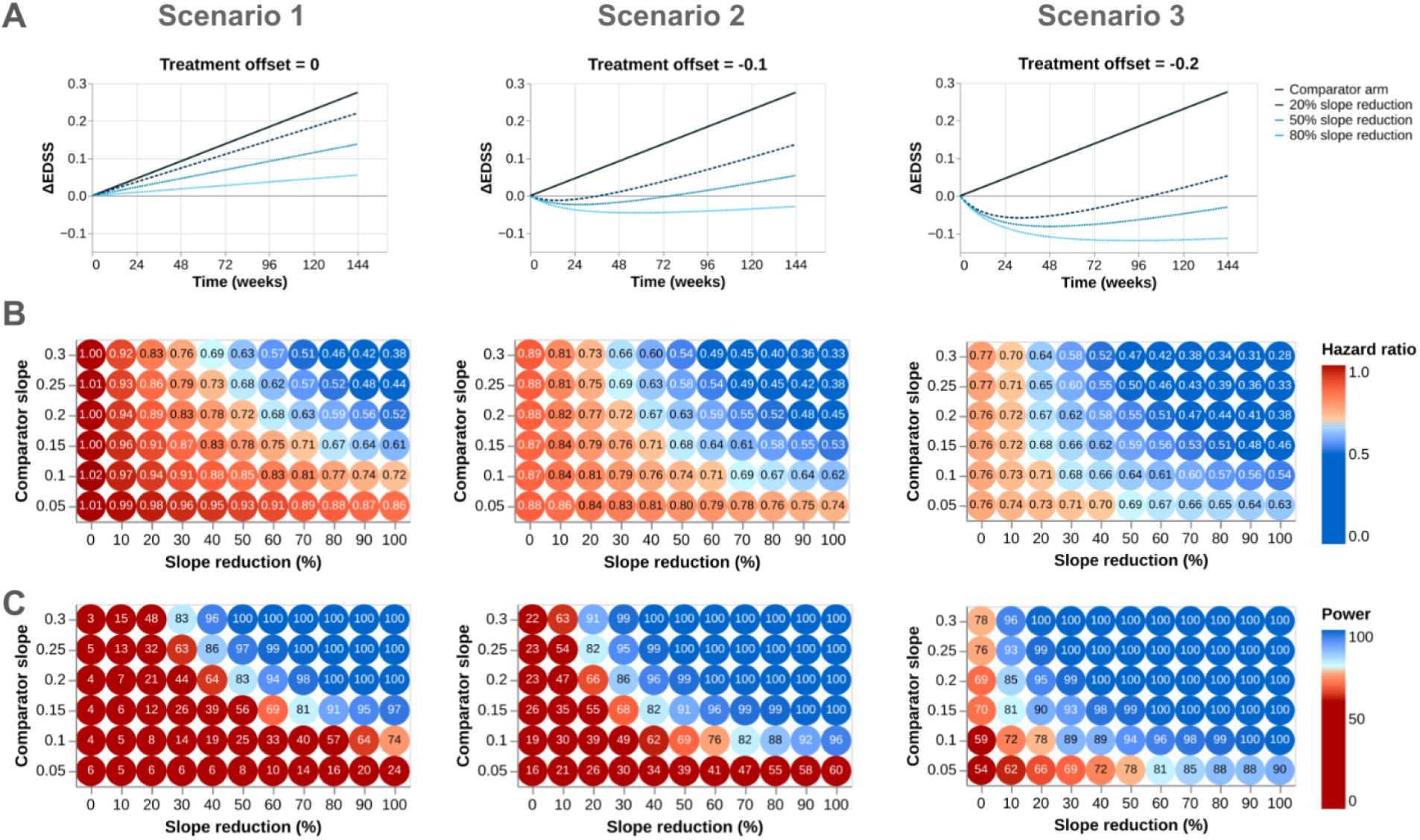
Simulations of different scenarios for new drug candidates targeting the remaining progression in pwMS treated with HEDMTs. Scenario 1, left: drug candidate with no fixed offset effect; scenario 2, center: drug candidate with a moderate fixed offset effect (-0.1); and scenario 3, right: drug candidate with a strong fixed offset effect (-0.2). As an illustration of each scenario, the comparator arm fixed progression is shown (for comparator slope = 0.1), as well as the fixed progression in the treatment arm assuming a 20%, 50% and 80% reduction of the comparator slope and the previously defined fixed offset effect (0, -0.1, and -0.2, respectively, panel A). For each scenario, different comparator slopes (y-axis on the heatmaps) and different drug effects on the slope were considered (x-axis on the heatmaps). For each comparator slope-drug effect pair, the average hazard ratio (panel B) and the power (i.e. the proportion of simulations with a CoxPH HR p-value < 0.05, panel C) were estimated (1,000 simulations). Except for the fixed slope and/or the fixed offset effect, all other parameters were similar between the two treatment arms, especially the interindividual variability of the slope and offset. Simulation parameters were chosen to represent typical phase III MS trials designs: 500 subjects per treatment arm, 144 weeks of follow-up per patient, and EDSS assessment every 12 weeks.

For the control arm, the fixed offset effect was set to 0 for the 3 scenarios, simulating a linear average progression as observed in the long term for pwMS treated with ocrelizumab (**Fig. 3A** comparator arm curve, in which a linear progression rate of 0.1 EDSS/year is shown). Regarding the treatment arm, the fixed offset effect was set as a constant in each scenario; 0 in scenario 1, -0.1 in scenario 2 (moderate offset effect, similar to what was observed for ocrelizumab in PPMS and SPMS), and -0.2 in scenario 3 (strong offset effect, similar to what was observed for ocrelizumab in RMS). Different progression patterns for the treatment arm (20%, 50%, and 80% slope reduction, and the scenario”s specific treatment offset effect), are shown in **Fig. 3A**.

Because the progression rate differs between different patient populations, we investigated different rates of comparator progression: 0.05, 0.10 (as shown in **Fig. 3A**), 0.15, 0.20, 0.25, and 0.30 EDSS/year (**Fig. 3B** and **C**, y-axis on the heatmaps). In addition to the offset, different relative reductions of the comparator slope were investigated within each scenario (**Fig. 3B** and **C**, x-axis on the heatmaps), ranging from no reduction (0%) to a complete stop, on average, of the progression (100%).

Within each scenario, we investigated how different pairs of comparator arm progression rate and drug effects would impact the HR and power of a typical phase III MS study (see Methods, Clinical Trials Simulations section for details). Briefly, EDSS change from baseline data was simulated at each timepoint (every 12 weeks for 144 weeks) for each subject using the model parameters previously estimated, without any covariate effect. From the simulated individual changes in EDSS, the time to first EDSS-CDP24 event was derived for each subject (500 per treatment arm). The resulting time-to-event data was used to fit a CoxPH model and estimate the HR between the comparator and the treatment arms. This process was repeated 1,000 times, to estimate the average HR (**Fig. 3B**) and the power (i.e. the proportion of the 1,000 simulations with a HR p-value < 0.05, **Fig. 3C**), for each combination of comparator slope (y-axis), treatment effect on the slope (x-axis), and treatment offset effect (scenarios 1, 2, and 3). The color-coding of the heatmaps was selected to represent typical targets for a phase III trial: a HR of 0.7, and 80% power. In addition, the corresponding average proportion of subjects with an event is shown in **Fig. S4**, and the proportion of simulations where the null hypothesis of constant hazard ratios was rejected (p<0.05) using the Schoenfeld residuals test is shown in **Fig. S5**.

These simulations allow us to understand how the comparator”s progression rate and the treatment effects collectively impact the time-to-event endpoint. First, we observe that the lower the comparator slope, the more difficult it is to reach low HR at constant relative drug effect on the slope. For example, in scenario 1, a reduction of the average slope by 40% with a comparator progression rate of 0.3 EDSS/year would result, on average, in a HR of 0.69 between the two arms. In the situation where the progression rate comparator slope is 0.1 EDSS/year, a similar relative reduction of the slope would result in an average HR of 0.88 (**Fig. 3B**, scenario 1). Consequently, detecting a statistically significant effect becomes more challenging in settings with a low comparator slope and no initial offset effect from the treatment; for the previous examples, the power drops from 96% in the first example to just 19% in the second (**Fig. 3C**, scenario 1).

The reasons for this power reduction are two fold. First, for a constant event rate in the comparator arm, a stronger treatment effect that results in a lower HR will increase statistical power (**Fig. 3C**, scenario 1). Second, even if two trial scenarios yield a similar HR, the scenario with a lower progression rate in the comparator arm will have less statistical power. This is because a slower progression rate results in fewer total disability events being observed during the study, given a fixed sample size and study duration (**Fig. S4**). For instance, in scenario 1, a 30% treatment effect with a comparator slope of 0.3 leads to an average HR of 0.76 and a power of 83%. In contrast, an 80% treatment effect on a comparator slope of 0.1 leads to similar HR (0.77), but a much lower power (57%).

As would be expected, the presence of an initial improvement from treatment (an offset effect) leads to a rapid separation of the mean change from baseline in EDSS between the two arms (**Fig. 3A**, scenario 2 and 3). On EDSS-CDP, this translates into lower event rates in the treatment arm, lower HRs, and higher statistical power (**Fig. 3B** and **C**, scenario 2 and 3, and **Fig.S4**). Interestingly, this does not lead to a significantly higher proportion of simulations violating the HR assumption (**Fig.S5)**.

## Discussion

In this study, we developed a mathematical model to investigate and quantify long-term longitudinal EDSS progression in pwMS treated with ocrelizumab. Using data from multiple phase III clinical trials, we characterized the average response to ocrelizumab by two components: 1) an initial decrease or stabilization of EDSS within the first months of treatment initiation (termed “offset effect”), followed by 2) a steady linear increase of EDSS.

The observed linearity of the long-term progression in pwMS treated with ocrelizumab is consistent with other reports of long-term disability progression in MS.^16,17^ Long-term progression rate is higher in progressive MS (**Fig. 2,A**), and is presumed to be the consequence of pathological mechanisms not fully addressed or controlled by ocrelizumab. Although dropouts could potentially impact the estimation of progression rates, a comparison of baseline characteristics between participants who dropped out and those who completed the trial at year 9 revealed no relevant differences (**Fig. S3B**), suggesting that dropouts do not have a relevant impact on these estimates. A delay in treatment effect, reported for IFN beta-1a or glatiramer acetate, was not considered in this work. There is no reported evidence of such an effect for ocrelizumab, and the observed linear average chronic progression (**Fig. 1B**) does not suggest such an effect.^18^

The observed initial decrease or stabilization of disability measured by EDSS, while less intuitive in a progressive disease such as MS, is consistent with previously reported findings, and with the rapid and sustained peripheral B-cell depletion observed with ocrelizumab. An analysis of the OPERA data showed that among subjects randomized to ocrelizumab, the proportion of subjects with EDSS disability improvement confirmed at 24 weeks (CDI24) after 2 years was 16.8%, considerably higher than the proportion of subjects with CDP24 (7.7%).^19^ A similar initial disability improvement was also observed for other lymphocyte-targeting drugs, such as ofatumumab, ublituximab, and alemtuzumab.^9,20–24^ This observation is commonly attributed to the profound, fast onset effect of these drugs on disease activity, which permits recovery from previous inflammation (and relapse) under conditions of a greatly attenuated inflammatory process, and consequent disability improvement on trial. In alignment with this observation, in our analysis, the presence of T1 Gd-enhancing lesions at baseline and a more inflammatory MS subtype was associated with a higher offset effect (**Fig. 2B,C**). Our formulation is that the offset effect observed in pwMS treated with ocrelizumab and other HEDMTs is mainly explained by resolving focal inflammation occurring shortly after treatment initiation.

In addition, we showed that while our model simulates longitudinal changes in EDSS, it is able to properly capture the standard event based clinical trial progression endpoint, EDSS-CDP24 (**Fig. 1C**). This finding allows us to investigate the relationship between changes in the magnitude of the offset effect or changes in the rate of long term progression, and the cumulative probability of EDSS-CDP24. This is of particular value when considering the design of future trials using event-based progression endpoints and highly effective comparators.

Our simulations highlight the key importance of the offset effect in time to disability progression event studies. Compared to a scenario with no offset effect (scenario 1), the presence of a moderate to strong offset effect (scenarios 2 and 3), delivers a much higher trial effect size (measured by HR) for similar treatment effects on long-term progression (**Fig. 3B**). This reveals a fundamental challenge to contemporary MS trial design: while the primary unmet need is slowing chronic disability, the current paradigm favors drugs that induce initial improvement over those effectively targeting the underlying progression rate. This bias is most evident when the progression rate in the comparator arm is low, which is commonly seen in pwMS treated with HEDMTs. Assuming a comparator arm progression rate similar to that observed in pwMS treated with ocrelizumab (0.02-0.12 EDSS/year, **Fig. 2A**), and with no offset in both treatment arms, the probability of observing a significant treatment effect on EDSS-CDP24 would be very low, even if the drug substantially reduces the progression rate (**Fig. 3B** and **C**, scenario 1).

The limited power of EDSS-CDP to detect meaningful treatment effects stems from its susceptibility to “random variation and measurement error”—as captured by the random effects in our model—and previously highlighted by Ebers *et al*.^*25*^ Furthermore, the dependency of EDSS-CDP (within acceptable trial durations) on the initial offset effect poses an additional significant barrier to the development of novel therapies targeting the underlying drivers of long-term disability accumulation.^4^ Promising neuroprotective therapies, that act by reducing axonal loss or glial inflammation, may be erroneously deemed ineffective if assessed as add-on therapies in time to event trial designs: without an associated offset effect to differentiate them from the comparator, their impact on the long-term disease course will remain statistically obscured.

We must acknowledge the limitations of our study, which include the reliance on EDSS data (a non-linear disability scale), the inclusion of clinical trial data of pwMS treated with ocrelizumab only, and the simulation assumptions. EDSS is a composite categorical scale with intervals between scores that do not represent equal increments of disability. This means that a particular EDSS progression rate (for example 1 point over 5 years) characterizes a variety of “true progression rates”, because the baseline EDSS influences its meaning: a shift from EDSS 3 to EDSS 4 represents a very different clinical scenario when compared to a shift from EDSS 6 to EDSS 7, and this influences the interpretation of the results. The response observed in pwMS treated in the ocrelizumab studies may not necessarily be generalizable to different patient populations, treatment interventions, and trial designs. It is also important to highlight the assumptions that were made for the clinical trial simulations, especially a similar interindividual variability for both treatment arms, which may not always hold true.

Our work enhances the understanding of disability progression in pwMS treated with HEDMTs, and the performance of EDSS-CDP in different scenarios. Specifically, our simulation results highlight the urgent need to develop the tools to design MS clinical trials differently, and evolve the design of those trials, in order to enable the development of new drugs targeting long-term disability progression in subjects with low or absent focal inflammation. In proof of concept studies this involves exploring and validating more sensitive disease measures that can directly capture gradually progressive pathology;^26–29^. In phase III studies, more granular disability assessments should be employed to increase signal, but will only offer meaningful improvements in the detection of an effect if they avoid additional measurement noise, and accurately reflect the lived disability of pwMS.^30^ Innovative trial designs represent an additional lever, including enrichment strategies for subjects at high risk of progression or the use of “time saved” analyses instead of time-to-event endpoints.^31^ It is crucial that future trials are properly powered to detect clinically meaningful effects on “real” disability progression, ensuring that we efficiently identify the next generation of MS therapies to address the remaining unmet need of pwMS.

## Author Contributions

*Concept and design:* Gaudry, Boareto.

*Acquisition, analysis, or interpretation of data:* All authors.

*Drafting of the manuscript:* Gaudry, Boareto.

*Critical review of the manuscript for important intellectual content:* All authors.

*Statistical analysis:* Gaudry, Thanei, Boareto.

*Supervision:* Bonati, Boareto.

## Acknowledgment

The authors thank all study participants, their families, the site staff, and the trials teams for their time and commitment to the trials. The authors also thank the following colleagues and MS experts for their critical feedback on method development and discussions: Dr. Sven Schippling, Dr. Cheikh Diack, and Dr. Frederik Buijs.

