## Supplementary Information for "Quantifying MS Progression in the Era of Highly Effective Therapy: Trial Design Implications"

#### Baseline Characteristics

**Supplementary Table 1:** Baseline characteristics of the included patients per MS subtype.

| | | PPMS | RMS | SPMS |
|---|---|---|---|---|
| **Subjects** | *n* | 743 | 763 | 353 |
| **Male** | *n* | 381 | 269 | 145 |
| | % | 51 | 35 | 41 |
| **Presence of T1 Gd+** | *n* | 156 | 314 | 71 |
| | % | 21 | 41 | 20.1 |
| **Age** | *mean* | 46.6 | 37.2 | 47.6 |
| | *sd* | 8.7 | 9.1 | 9.6 |
| **EDSS** | *mean* | 4.7 | 2.8 | 5.3 |
| | *sd* | 1.3 | 1.3 | 1.2 |
| | *median* | 4.5 | 2.5 | 6 |
| **Years since MS onset** | *mean* | 6.9 | 6.7 | 16.2 |
| | *sd* | 4.3 | 6.2 | 8.5 |
| **BMI** | *mean* | 25.3 | 26.1 | 24.8 |
| | *sd* | 5.0 | 5.8 | 4.8 |
| **Follow-up duration in years** | *mean* | 5.9 | 7.7 | 3.0 |
| | *sd* | 3.4 | 3.2 | 0.9 |
| | *median* | 3.8 | 9.5 | 3.3 |

#### Mathematical Model

A Bayesian hierarchical model was employed to analyze the relationship between change in EDSS, time, and subject-specific characteristics. The model was implemented in Stan and is described below.

The core of the model is a non-linear function describing the change from baseline in EDSS over time for each subject using two additive components: offset effect and linear progression.

Specifically, for the observation from a given subject $s$ at time $t$, the estimated EDSS change, $\hat{y}_{s,t}$, is modeled as:

$$\hat{y}_{s,t} = \overbrace{\omega_s \cdot \frac{t}{\tau_{50} + t}}^{\text{Offset effect}} + \underbrace{\gamma_s \cdot t}_{\text{linear progression}} \tag{1}$$

where $\omega_s$ refers to the subject $s$ maximal offset effect. The parameter $\tau_{50}$ is a global parameter representing the time at which half of the maximum offset is reached, and it is assumed to be common across all subjects. $\gamma_s$ represent the long term progression rate of subject $s$.

The individual parameters $\omega_s$ and $\gamma_s$ are further modeled hierarchically to account for population-level trends and individual deviations:

$$\omega_s = \mu_\omega + X_s \cdot \beta_1 + u_{1,s} \tag{2}$$

$$\gamma_s = \mu_\gamma + X_s \cdot \beta_2 + u_{2,s} \tag{3}$$

Here, $\mu_\omega$ is the fixed population-level maximum offset, and $\mu_\gamma$ is the fixed population-level progression rate. $X_s$ is the vector of $d$ covariates for subject $s$. $\beta_1$ and $\beta_2$ are vectors of $d$ coefficients representing the effects of covariates on the offset and slope, respectively. The terms $u_{1,s}$ and $u_{2,s}$ are subject-specific random effects for the offset and slope, respectively. These random effects are assumed to follow a double exponential (Laplace) distribution centered at zero:

$$u_{1,s} \sim Laplace(0, \sigma_\omega) \tag{4}$$

$$u_{2,s} \sim Laplace(0, \sigma_\gamma) \tag{5}$$

Finally, the observed EDSS change values $y_{s,t}$ are assumed to be normally distributed around their predicted values $\hat{y}_{s,t}$ with a residual error $\sigma_e$:

$$y_{s,t} \sim N(\hat{y}_{s,t}, \sigma_e) \tag{6}$$

### Prior Distributions

Prior distributions were chosen to regularize the model and incorporate existing knowledge.

$$\begin{gathered} \beta \sim N(0, 0.5) \\ \mu_\omega \sim N(0, 0.5) \\ \mu_\gamma \sim N(0, 0.5) \\ \tau_{50} \sim N(1, 5) \text{ truncated at 0 to ensure positivity} \end{gathered} \tag{7}$$

All other model parameters were assigned uniform prior distributions.

#### Model Implementation

The model was implemented using Stan (Carpenter et al., 2017), a probabilistic programming language for Bayesian inference. Markov Chain Monte Carlo (MCMC) simulations were used to sample from the posterior distributions of the model parameters. The number of chains was set to 4, with 1,000 warm-up iterations, and 1,000 sampling iterations.

#### Convergence Diagnostics

Convergence diagnostics were thoroughly assessed. Treedepth for all transitions was satisfactory. No divergent transitions were observed during sampler operation. The E-BFMI (energy-Bayesian fraction of missing information) for Hamiltonian Monte Carlo potential energy was satisfactory, indicating efficient sampling. Effective sample size (ESS) values were sufficient, and split R-hat values for all parameters were satisfactory, suggesting good chain mixing and convergence. Summary of the posterior distribution, ESS and R-hat values of the main parameters are available in Supplementary Table 2.

**Supplementary Table 2:** Summary of the posterior distributions, ESS, and R-hat for the model's main parameters.

| | mean | sd | HDI 2.5% | HDI 97.5% | ESS | R-hat |
|---|---|---|---|---|---|---|
| $\mu_\omega$ | -0.219 | 0.042 | -0.304 | -0.140 | 1054 | 1.001 |
| $\mu_\gamma$ | 0.045 | 0.0084 | 0.029 | 0.062 | 889 | 1.003 |
| $\sigma_\omega$ | 0.648 | 0.020 | 0.608 | 0.687 | 1421 | 1.000 |
| $\sigma_\gamma$ | 0.118 | 0.004 | 0.110 | 0.125 | 2306 | 1.002 |
| $\tau_{50}$ | 0.530 | 0.027 | 0.480 | 0.585 | 687 | 1.001 |
| $\sigma_e$ | 0.463 | 0.002 | 0.460 | 0.467 | 5044 | 1.001 |

#### Simulation Model

The previously developed non-linear mixed-effects model was used as a simulation tool to evaluate different scenarios of drug development in MS. For each scenario, the fixed slope and offset of each arm, control (1) and treatment (2), were set to defined values. For each set of parameters, 1,000 clinical trials were simulated assuming: a 1:1 randomization between treatment arms, an EDSS assesment every 12 weeks for 144 weeks, 1,000 subjects in total, and no dropout. The core of the model is the same non-linear function describing the change in EDSS over time for each subject using two additive components, offset effect and linear progression (same components that were used to fit the data), without any covariates effect on these parameters. as we assume covariates effects would depend on the trial design and treatments' modes of action.

Specifically, the mean EDSS change for a given subject $s$ at time $t$ in treatment arm $a$, with a ∈ $\{1, 2\}$, is defined by the following equation as:

$$\hat{y}_{s,t,a} = \overbrace{\omega_s \cdot \frac{t}{\tau_{50} + t}}^{\text{Offset effect}} + \underbrace{\gamma_s \cdot t}_{\text{linear progression}} \tag{8}$$

where $\omega_s$ refers to the subject $s$ maximal offset effect. The parameter $\tau_{50}$ is a global parameter representing the time at which half of the maximum offset is reached. $\gamma_s$ represent the long term progression rate of subject $s$. The individual parameters $\omega_s$ and $\gamma_s$ are further defined as follows:

$$\omega_s = \mu_{\omega,a} + u_{1,s} \tag{9}$$

$$\gamma_s = \mu_{\gamma,a} + u_{2,s} \tag{10}$$

Here, $\mu_{\omega,a}$ is the fixed maximum offset for arm $a$, and $\mu_{\gamma,a}$ is the fixed progression rate for arm $a$. These 2 parameters are defined according to the simulations scenario. The terms $u_{1,s}$ and $u_{2,s}$ are subject-specific random effects for the offset and slope, respectively. These random effects are assumed to follow a double exponential (Laplace) distribution centered at zero with scale $\sigma_\omega$ and $\sigma_\gamma$, respectively, identical for all subjects in the simulated trial:

$$u_{1,s} \sim Laplace(0, \sigma_\omega) \tag{11}$$

$$u_{2,s} \sim Laplace(0, \sigma_\gamma) \tag{12}$$

Finally, the simulated EDSS change values $y_{s,t}$ are assumed to be normally distributed around their predicted values $\hat{y}_{s,t}$ with a residual error $\sigma_e$:

$$y_{s,t,a} \sim N(\hat{y}_{s,t}, \sigma_e) \tag{13}$$

For each simulated trial, the following parameters $\tau_{50}$, $\sigma_\omega$, $\sigma_\gamma$, and $\sigma_e$, identical for all subjects, are drawn from normal distributions with location and scale corresponding to the posterior distributions estimated from the data at the previous step (see Mathematical Model).

The predicted EDSS changes from baseline were rounded to the nearest half-integer, and each subject was assigned a baseline EDSS by sampling with replacement from the dataset of PPMS subjects included in the study. The time to EDSS-CDP24 event, as defined in the main body of the article, was then derived for each subject of the simulated trial.

#### Supplementary Figures

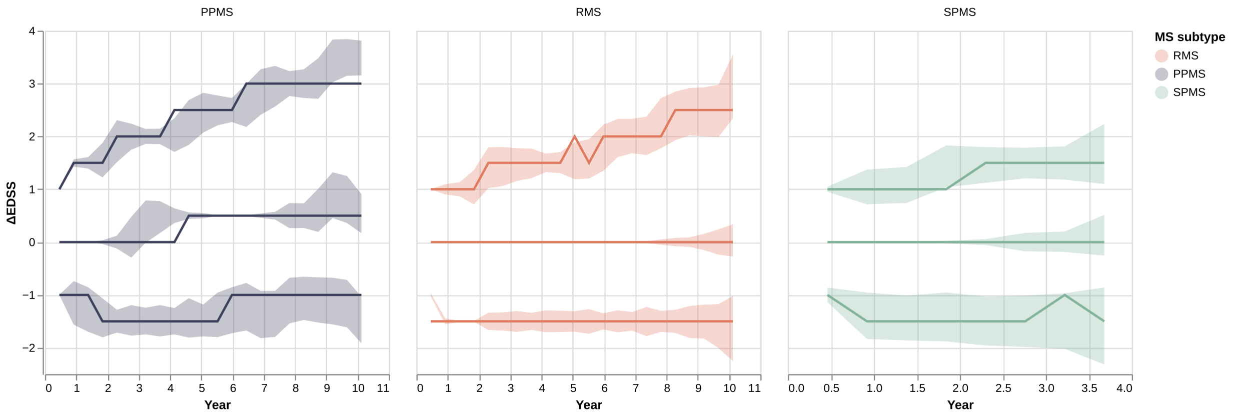


**Figure S1**: Percentiles (5, 50 and 95) in observed data (line) and confidence interval from model predictions (shaded area), per MS subtype.

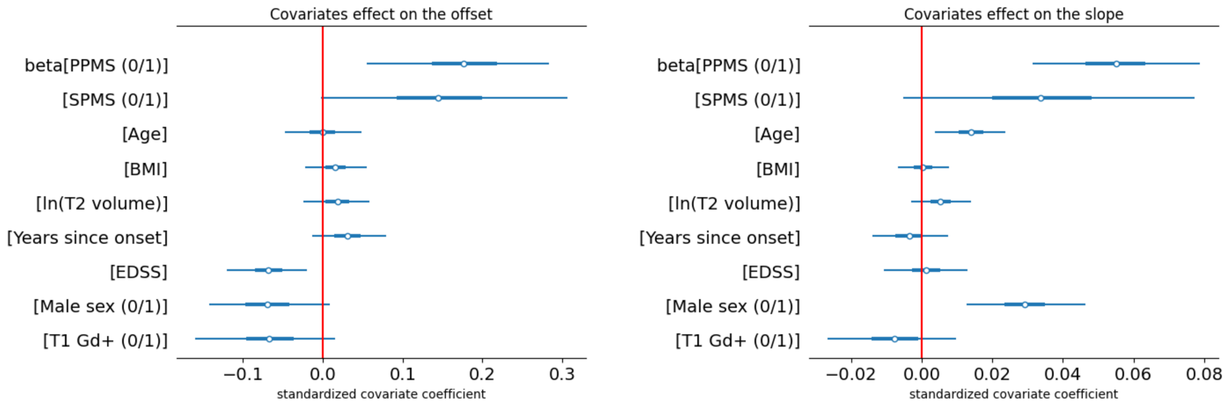


**Figure S2**: Covariates coefficient (β) on the offset (left) and slope (right) parameters (median and 95% credible intervals).

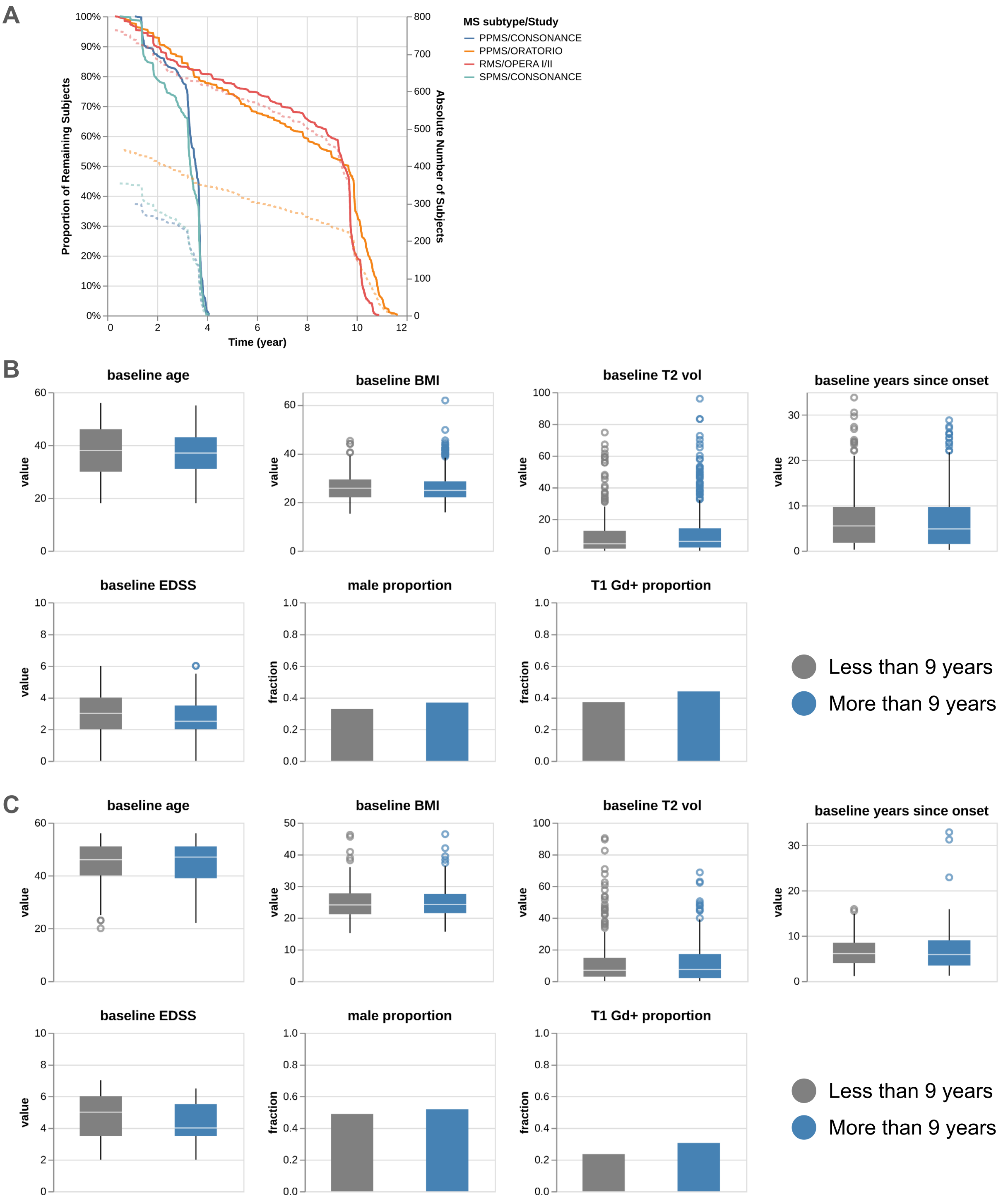


**Figure S3**: Proportion (solid line) and absolute numbers (dashed line) of included subjects as a function of time for PPMS/ORATORIO, PPMS/CONSONANCE, SPMS/CONSONANCE, and RMS/OPERA I/II (A). Baseline chracteristics for subjects who dropped out before (grey) and after (blue) 9 years of follow-up, respectively, for RMS (B) and PPMS (C).

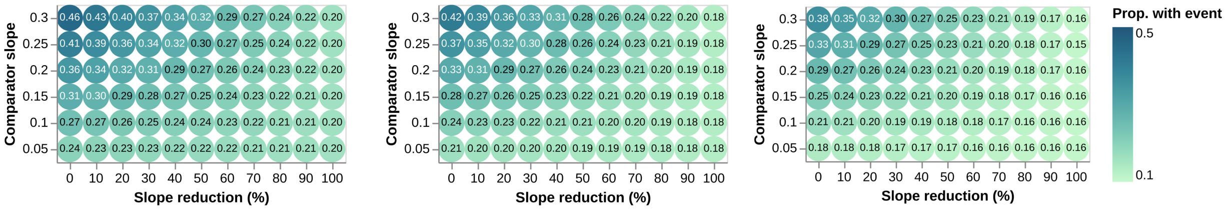


**Figure S4**: Average proportion of patients with an EDSS-CDP24 event in the treatment arm, for scenarios 1 (left), 2 (center), and 3 (right).

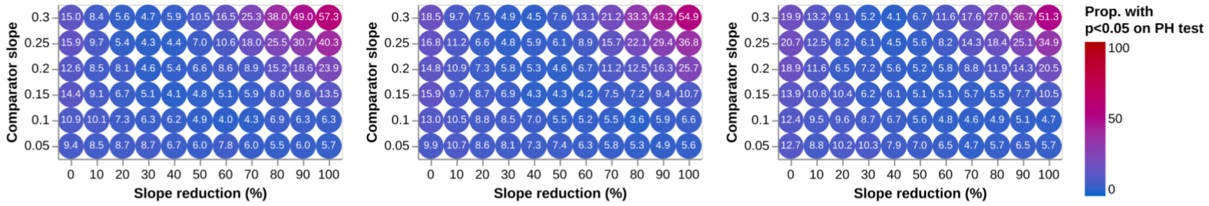


**Figure S5**: Proportion of simulations where the null hypothesis of constant hazard ratios was rejected (p<0.05) using the Schoenfeld residuals test (*lifelines proportional_hazard_test()* function, with rank-transformed time). Results indicate the frequency of PH assumption violations between the two treatment arms for Scenario 1 (left), Scenario 2 (center), and Scenario 3 (right).
